# Construction of a short version of the Montreal Cognitive Assessment (MoCA) rating scale using Partial Least Squares analysis

**DOI:** 10.1101/2022.05.05.22274726

**Authors:** Solaphat Hemrungrojn, Arisara Amrapala, Michael Maes

## Abstract

**Background:** The Montreal Cognitive Assessment (MoCA) rating scale is frequently used to assess cognitive impairments in amnestic mild cognitive impairment (aMCI) and Alzheimer’s disease (AD).

**Objectives:** The aims of this study are to a) evaluate the construct validity of the MoCA and its subdomains or whether the MoCA can be improved by feature reduction, and b) develop a short version of the MoCA (MoCA-Brief).

**Methods:** We recruited 181 participants, divided into 60 healthy controls, 61 aMCI, and 60 AD patients.

**Results:** The construct reliability of the original MoCA was not optimal and could be improved by deleting one subdomain (Naming) and five items, namely Clock Circle, Lion, Digit Forward, Repeat 2^nd^ Sentence, and Place, which showed inadequate loadings on the extracted latent vectors. To construct the MoCA-Brief, the reduced model underwent further reduction and feature selection based on model quality data of the outer models. We produced a MoCA-Brief rating scale comprising five items, namely Clock Time, Subtract 7, Fluency, Month, and Year. The first latent vector extracted from these five indicators showed adequate construct validity with an Average Variance Extracted of 0.599, composite reliability of 0.822, Cronbach’s alpha of 0.832 and rho_A of 0.833. The MoCA-Brief factor score showed a strong correlation with the total MoCA score (r=0.98, p<0.001) and shows adequate concurrent, test-retest, and inter-rater validity.

**Conclusion:** The construct validity of the MoCA may be improved by deleting five items. The new MoCA-Brief rating scale deserves validation in independent samples and especially in other countries.

## 1. Introduction

The proportion of the world’s elderly population (age over 60 years) is projected to increase to 22% by 2050 (almost double the proportion in 2015) (Kanasi, Ayilavarapu and Jones, 2016). With this, the prevalence of dementia is also expected to significantly increase (Norton, Matthews and Brayne, 2013). Accounting for 60-80% of all dementia cases is Alzheimer’s Disease (AD) (Alzheimer’s Association, 2020), a progressive brain disorder that is a major global healthcare burden. This neurodegenerative disease is clinically characterized by a gradual decline in cognitive ability, which is in part due to the presence of neuritic beta-amyloid plaques and neurofibrillary tau protein tangles (Murphy and LeVine, 2010).

Another condition characterized by neurocognitive decline is Mild Cognitive Impairment (MCI). MCI is an intermediate stage that is often, but not always, a transitional phase between normal age-related cognitive decline and dementia (Anderson, 2019). As defined by Petersen *et al*. (1997), the diagnosis for MCI is made when a subject: (1) has memory complaints and abnormal memory function for their age, (2) has normal general cognitive function and activities of daily living, and (3) does not have dementia. MCI can be further divided into non-amnestic (naMCI) and amnestic (aMCI) MCI that can either be multiple domain or single domain. Those with single-domain aMCI are at higher risk of developing dementia, whereas naMCI is associated with non-AD dementias (Csukly *et al*., 2016).

A well-known tool for the detection of AD and MCI is the Montreal Cognitive Assessment (MoCA). The MoCA was developed by Nasreddine *et al*. (2005) to address shortcomings of the Mini-Mental State Examination (MMSE), a widely used cognitive function test that was less sensitive to MCI. The MoCA is a 10-minute test for older adults that evaluates cognitive domains such as executive function, visuospatial function, memory, language, orientation, and attention. Since its appearance, many validation studies continuously report that the MoCA (English version and other language versions) had good diagnostic accuracy in discriminating MCI and AD patients, and better psychometric properties compared to other cognitive impairment tests (Fujiwara *et al*., 2010; Freitas, Simões, Alves, *et al*., 2012; Freitas *et al*., 2013; Azdad *et al*., 2019; Freud *et al*., 2020; Serrano *et al*., 2020; Hemrungrojn *et al*., 2021).

Nevertheless, regarding the construct of the MoCA test itself, the psychometric properties of the test remain under debate as not many studies have delineated its factorial structure and measurement invariance. The MoCA’s psychometric properties are an important aspect to analyze as it is concerned with the reliability of the total score in representing the subject’s real cognitive status as well as the interpretations drawn from it. For the total MoCA score to be considered viable and true, the factorial structure of the test should be unidimensional (i.e., have one general factor present that relates all the items together by explaining more than 50% of the variance) regardless of how many other dimensions the model has. Without a general factor present, the total score will not represent the overlying construct (the subject’s cognitive status) and will therefore be less meaningful (Sala *et al*., 2020). Studies that have explored the MoCA’s construct have yielded different, sometimes contrasting results on aspects such as the unidimensionality of the MoCA and the number of factors.

The original authors of the MoCA presumed the unidimensionality of the test and proposed a six-dimensional factorial model (Hobson, 2015). Unidimensionality has also been assumed in other studies, albeit with a different number of factors amongst the studies, in various contexts such as its use with geriatric (Koski, Xie and Finch, 2009; Sala *et al*., 2020), MCI and AD (Freitas, Simões, Marôco, *et al*., 2012; Abdul Karim and Venkatachalam, 2021), early-stage Parkinson’s Disease (PD) (Kletzel, Louise-Bender Pape and Mallinson, 2016), and cancer patients (Arcuri *et al*., 2015).

Contrastingly, other studies found multidimensionality with no clear general factor. For example, Coen *et al*. (2015) reported that even though the MoCA had an adequate model fit for the six-factor model using confirmatory factor analysis (CFA), exploratory factor analysis (EFA) showed no clear patterns in item cross-loading for both a three- and six-factor models. The authors concluded that the MoCA was not a suitable in-depth neuropsychological assessment for domain-specific evaluation. Another example is the use of MoCA in PD patients whereby a six-factor model was found using EFA, however, the validity of the model was not supported by CFA (Smith *et al*., 2020).

Apart from unidimensionality, other measures such as construct validity and internal consistency help determine the strength of the tool. Construct validity measures whether the tool in question is strongly associated with other similar cognitive tests, whilst internal consistency measures whether the items that make up the tool in question are closely related. The MoCA has generally demonstrated good construct validity (average variance extracted, AVE, of 0.46 – 0.75) (Freitas, Simões, Marôco, *et al*., 2012) and good internal consistency (Cronbach’s alpha = 0.83 - 0.903) for MCI and AD patients (Nasreddine *et al*., 2005; Freitas, Simões, Marôco, *et al*., 2012; Hemrungrojn *et al*., 2021). These measures have also been reported to be good, however lower, in other populations such as healthy older adults and those with major depressive disorder (Cronbach’s alpha = 0.60 - 0.64) (Cooley *et al*., 2015; Srisurapanont *et al*., 2017). Collectively, the presence of an underlying unidimensionality, the number of factors, the construct validity, and the reliability of the MoCA requires further exploration to ensure its accuracy.

Despite the MoCA being a fast-screening test that requires only 10-15 minutes to complete (Mast and Gerstenecker, 2010), this time frame remains too long as (1) the elderly population may struggle to complete the full MoCA version, and (2) the full version requires a physician or healthcare profession to administer it to patients, which may interfere with their healthcare routine and/or make it difficult to effectively screen patients due to staff shortages. Accordingly, a short form of the MoCA to address its current constraints is needed. Even though multiple different short MoCA versions have been developed (Liew, 2019), these forms had limited generalisability. Hence, it is essential to develop a validated short Thai MoCA version for the Thai population.

The aims of the present study were to a) evaluate the validity of the MoCA subdomains and whether the total score is an accurate representation of cognitive status or whether the MoCA can be improved by feature reduction, and b) develop a short version of the MoCA (MoCA-Brief).

## 2. Methods

### 2.1 Participants

Data from our previous study (Hemrungrojn *et al*., 2021) was utilized in this analysis. Of the total 181 participants, 60 were healthy controls (HC), 61 were individuals with aMCI, and 60 were AD patients from King Chulalongkorn Memorial Hospital, Chao Phraya Abhaibhubejhr Hospital, and Angthong Hospital, Thailand. All the participants were examined from March 2012 to June 2015. Geriatric psychiatrists and senior neurologists diagnosed and assessed all study participants. They also collected clinical history and performed neurological and physical examinations on the patients. AD patients in this study were diagnosed using the National Institute of Neurological and Communication Disorders and Stroke/AD and Related Disorders Association (NINCDS-ADRDA) criteria (Blacker *et al*., 1994), Clinical Dementia Rating (CDR) scale score of 1-2, and impaired activities of daily living. For aMCI patients, the diagnosis was made using Petersen’s criteria and CDR of 0.5 (a score of 0.5 in the memory subdomain is also required). HCs had a CDR of 0 and subjective memory complaints were absent. Patients and controls were excluded if they had: (1) other neurological diseases (e.g. PD, epilepsy, traumatic brain injury, multiple sclerosis, encephalitis, and meningitis), (2) other dementia syndromes (e.g. vascular dementia and frontotemporal dementia), (3) medical diseases (e.g. chronic obstructive pulmonary disease, chronic kidney disease, severe heart disease, uncontrolled hypothyroidism, cancer, and vitamin B12 deficiency), (4) axis I psychiatric disorders (e.g. schizophrenia, major depressive disorder, and substance abuse), and (5) abnormal thyroid, vitamin B12, liver, and kidney laboratory tests.

### 2.2 Clinical Dementia Rating (CDR) Scale

The CDR is a global rating scale for identifying overall dementia severity (Morris, 1993). This global rating score is obtained from individually rating a subject’s memory, judgment and problem solving, orientation, home and hobbies, community affairs, and personal care at a 0- to 5-point scale, with 0 meaning “absent” and 5 meaning “terminal.”

### 2.3 Montreal Cognitive Assessment (MoCA)

The translated and validated Thai version of the MoCA (MoCA-Thai) was used (Hemrungrojn *et al*., 2021). The items of the test are divided into 7 subdomains: (1) Visuospatial/Executive (modified trail-making B task: 1 point, clock drawing: 3 points, and cube copying: 1 point), (2) Naming (lion, rhinoceros, and camel: 3 points), (3) Attention (digit forward and backward: 2 points, target detection using tapping: 1 point, and serial subtraction: 3 points), (4) Language (sentence repetition: 2 points, and phonemic fluency: 1 point), (5) Abstraction (2-item verbal abstraction: 2 points), (6) Delayed Recall (recall of 5 nouns after 5 minutes: 5 points), and (7) Orientation (orientation to time and place: 6 points). The total sum of all individual scores (out of 30 maximum possible points) is said to represent the severity of cognitive impairment. The MoCA-Thai had high test-retest reliability (*r* = 0.990, *p* < 0.01, and Cronbach’s alpha = 0.923) and inter-rater reliability (Kappa = 0.994 at *p* < 0.01).

It is generally mentioned in the literature that the MoCA evaluates six cognitive domains. Nevertheless, the items on the test sheet are divided into seven domains: (1) Visuospatial/Executive, (2) Naming, (3) Attention, (4) Language, (5) Abstraction, (6) Delayed Recall, and (7) Orientation. For this seven-domain construct, the ‘Visuospatial’ and ‘Executive’ functions from the six-domain form are combined into one, the ‘Memory’ domain is renamed to ‘Delayed Recall’, and ‘Naming’ and ‘Abstraction’ domains are added. The differences in the MoCA subdomain divisions seem to be trivial as the decision depends upon how and what the respective researcher intends on analysing.

### 2.4 Mini-Mental State Examination (MMSE)

The translated and validated Thai version of the MMSE (TMSE) was used (Train the Brain Forum Committee, 1993) in this study. The test was developed to measure cognitive function amongst the elderly. Test items assess orientation, memory, language, visuospatial skills, and attention, and is scored out of a total of 30 points (Folstein, Folstein and McHugh, 1975). Different cutoff scores for the TMSE/MMSE are used depending on the level of education.

### 2.5 Statistical analysis

One-way analysis of variance (ANOVA) or the Kruskal-Wallis test was used to compare scale variables across groups, while category variables were evaluated using analysis of contingency tables (χ^2^ tests). Correlations between variables were checked with Spearman’s rank-order correlation coefficients). Neural network analysis was performed using AD and controls as the output variables and MoCA items as the input variables. We used an automated feed-forward architecture, multilayer perceptron neural network model and trained the model using two hidden layers, each with up to four nodes and 20-50 epochs using minibatch training with gradient descent. As a stopping rule, a single consecutive step with no further decline in the error term was chosen. We retrieved three samples: a) a training sample (47.7 percent) to estimate the network’s parameters, b) a holdout sample (33.3 percent) to verify the final network’s accuracy, and c) a testing sample (20.0 percent) to avoid overtraining. We calculated error, relative error, and the magnitude and relative magnitude of all input variables. All statistical analyses were conducted using IBM SPSS windows version 28.

Feature selection and reduction were performed based on PLS analysis (SmartPLS 3) (Ringle, Wende and Becker, 2015). Each variable was input as a single indicator or as a latent vector using reflective models. The quality criteria of the outer models were checked by assessing the construct and convergence reliability of the outer models whereby composite reliability should be > 0.75, Cronbach’s alpha coefficient > 0.7, rho A > 0.7 and AVE > 0.5, and all loadings on the latent vectors should be > 0.65 at *p* < 0.001. Confirmatory Tetrad Analysis helped confirm that the models were not misspecified as reflective models. PLS analysis was performed using 5,000 bootstraps. The quality of the final PLS model is checked using the standardized root mean square residual (SRMR) which should be <0.08.

## 3. Results

### 3.1 Sociodemographic and clinical data

**Table 1** shows that there were no significant differences in sex ratio between the study samples while there were intergroup differences in age, education and income. Generally, controls and aMCI individuals had significantly higher years of education and income than AD patients and mean age increased from controls to aMCI to AD. All scores, either total TMSE and MoCA and the MoCA subdomain scores (except Naming) were significantly different between the three groups and decreased from controls to aMCI to AD. The Naming subdomain was significantly lower in AD than in the other 2 groups.

**Table 1.** Sociodemographic and clinical features of healthy controls (HC), amnestic mild cognitive impairment (aMCI), and Alzheimer’s Disease (AD)

| Variables | Participants (Total = 181) | | | $F^s / \chi^2$ | *p* value |
| --- | --- | --- | --- | --- | --- |
|  | HC (*n* = 60)<sup>A</sup> | aMCI (*n* = 61)<sup>B</sup> | AD (*n* = 60)<sup>C</sup> |  |  |
| Age, years* | 67.9 (± 6.4) | 72.1 (± 7.0) | 76.8 (± 8.0) | 23.39 | < 0.000 |
| Sex, M/F | 21/39 | 22/39 | 19/42 | 0.363 | 0.834 |
| Education, years | 11.4 (± 5.4)<sup>C</sup> | 10.0 (± 5.5)<sup>C</sup> | 7.1 (± 5.1)<sup>A,B</sup> | 10.20 | < 0.000 |
| Income, baht/month | 15,795 (± 22,236)<sup>C</sup> | 12,789 (± 29,200)<sup>C</sup> | 3,783 (± 7888)<sup>A,B</sup> | 4.78 | 0.010 |
| Total TMSE Score* | 28.2 (± 1.8) | 26.2 (± 2.2) | 19.7 (± 4.8) | 115.60 | < 0.001 |
| VS/E* | 4.3 (1.1) | 3.4 (1.4) | 1.3 (1.2) | 94.04 | <0.001 |
| Naming | 3.0 (0.3)<sup>C</sup> | 2.7 (0.6)<sup>C</sup> | 2.0 (0.9)<sup>A,B</sup> | 32.11 | <0.001 |
| Attention* | 5.5 (0.9) | 4.9 (1.2) | 3.2 (1.9) | 45.07 | <0.001 |
| Language* | 2.0 (1.2) | 1.4 (1.0) | 0.3 (0.6) | 44.29 | <0.001 |
| Abstract* | 1.3 (0.9) | 0.9 (0.8) | 0.3 (0.5) | 26.58 | <0.001 |
| Recall* | 3.0 (1.7) | 1.5 (1.4) | 0.2 (0.6) | 64.49 | <0.001 |
| **Orientation\*** | 5.9 (0.3) | 5.6 (0.7) | 3.0 (1.6) | 139.56 | <0.001 |
| **Total MoCA score\*** | 25.2 (3.0) | 20.9 (3.9) | 11.1 (4.6) | 206.63 | <0.001 |
| **MoCA-Brief factor score\*** | 0.744 (0.366) | 0.390 (0.626) | -1.110 (0.727) | 165.38 | <0.001 |

Data is shown as mean (SD) or as a ratio. <sup>\$</sup>All df = 2/179, except sex (df = 2); F: results of ANOVA and $\chi^2$ : results of contingency analysis; <sup>A-C</sup>: Pairwise comparisons among group means; \*All significantly different between the three study groups (posthoc comparisons at $p < 0.001$ )
TMSE: Thai Mini-Mental State Examination; VS/E: Visuospatial/Executive subdomain; MoCA: Montreal Cognitive Assessment.

### 3.2 Initial PLS Analysis with All MoCA Items in the seven subdomains

Figure 1. shows a first model based on the scores on the seven subdomains which were entered as indicators of a latent vector, dubbed the “seven-domain MoCA” latent vector (LV). The latter LV was connected with all latent vectors extracted from all items of the seven subdomain scores, dubbed the Visuospatial/Executive (VS/E), Naming, Attention, Language, Abstract, Delayed Recall, and Orientation LVs. Fluency was entered as the total scores of the fluency test (Fluency 1) and the dichotomized (≥ 11 is one point, Fluency 2) values. PLS analysis (5000 bootstraps, factor weighting scheme with maximum 1000 alterations and 7 as stop criterion) shows that all seven subdomain scores loaded significantly (all *p* < 0.001) and sufficiently (all > 0.65) on an LV with an AVE = 0.551, composite reliability = 0.895, Cronbach’s alpha = 0.862, and rho_A = 0.870. Most subdomain LVs showed adequate AVE (> 0.517), composite reliability (> 0.786), Cronbach alpha (> 0.806) and rho_A (> 0.769), although Cronbach’s alpha for Naming (0.593) and Attention (0.691), and the rho_A for Naming (0.634) were insufficient. These results show that the construct reliability of Naming is less than adequate and that this domain may be deleted from the model. Moreover, some item loadings were too low, namely Clock Circle on the VS/E LV, Lion (n1) on the Naming LV, Digit Forward (DF) on the Attention LV, Repeat 2^nd^ Sentence (repeat 2) on the Language LV, and Place on the Orientation LV.

**Figure 1.**
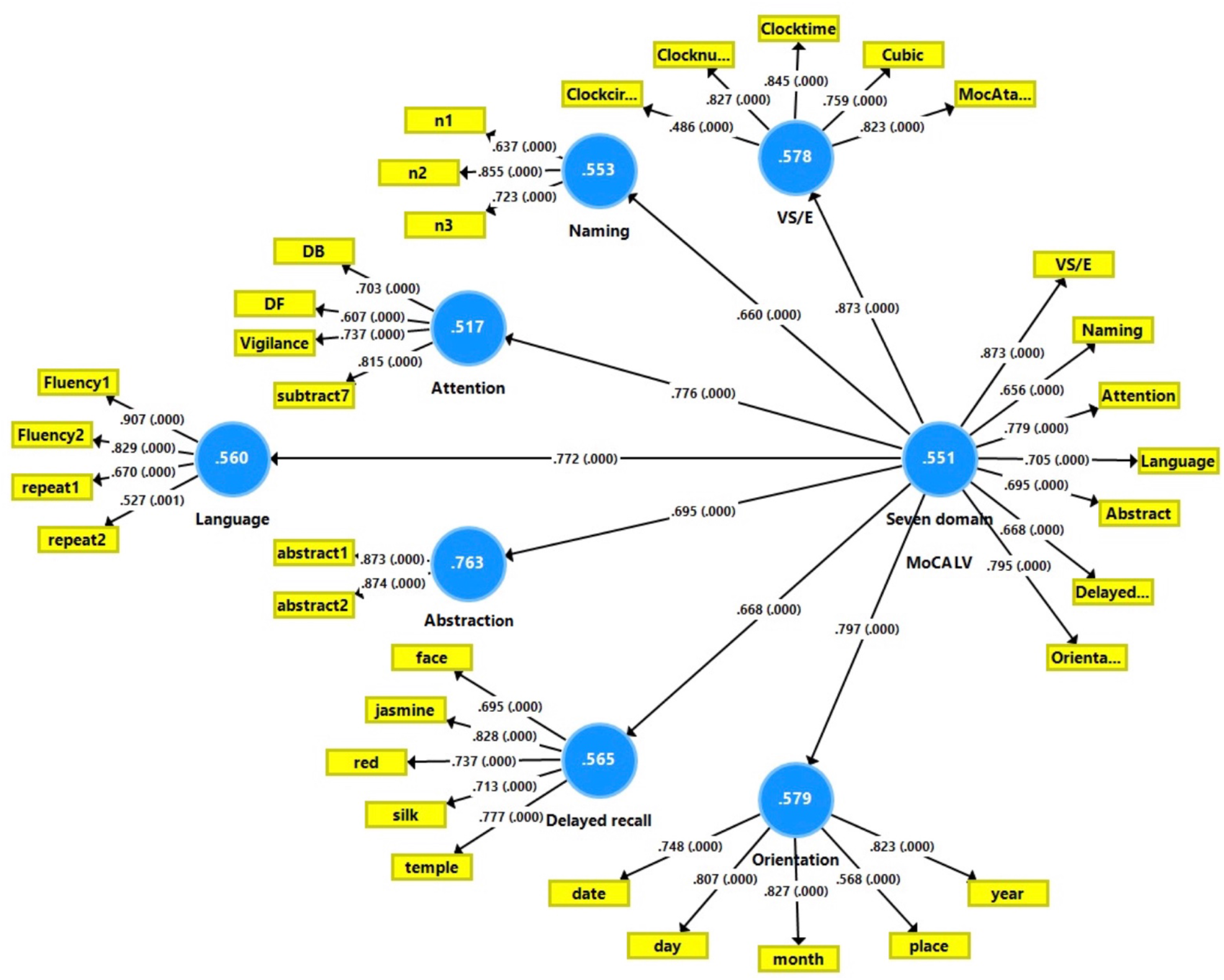
A first Partial Least Squares (PLS) model based on the scores on the seven subdomains, which were entered as indicators of a latent vector, dubbed the “seven-domain MoCA” latent vector (LV). This latter LV was connected with all LVs extracted from all items of the seven subdomain scores, dubbed the Visuospatial/Executive (VS/E), Naming, Attention, Language, Abstraction, Delayed Recall, and Orientation LVs. Shown are the loadings (with *p* values) for the outer models and path coefficients (exact *p* values) for the inner model are shown. White figures in blue circles denote Average Variance Extracted (AVE).

### 3.3. The reduced model shows improved performance

The reduced model, shown in **Figure 2**, was produced after feature reduction by deleting the Naming subdomain and the aforementioned 5 items from the analysis. This model shows a better performance in terms of construct convergence and reliability validity. All six subdomain scores loaded significantly (all *p* < 0.001) and sufficiently (all > 0.681) on a LV with an AVE = 0.583, composite reliability = 0.893, Cronbach’s alpha = 0.855, and rho_A = 0.863. All six subdomain LVs showed adequate AVE (> 0.565), composite reliability (> 0.814), Cronbach’s alpha (> 0.770), and rho_A (> 0.804). Nevertheless, it is notable that Attention performed worse with a rho_A = 0.675 and Cronbach’s alpha = 0.658. As such, the 21-item construct shown in **Figure 2** could be used to compute the summed scores on the six subdomains, and the LV or the sum of the latter may be used to reflect overall cognitive abilities. Psychometrically, this reduced model is more adequate than the original MoCA.

**Figure 2.**
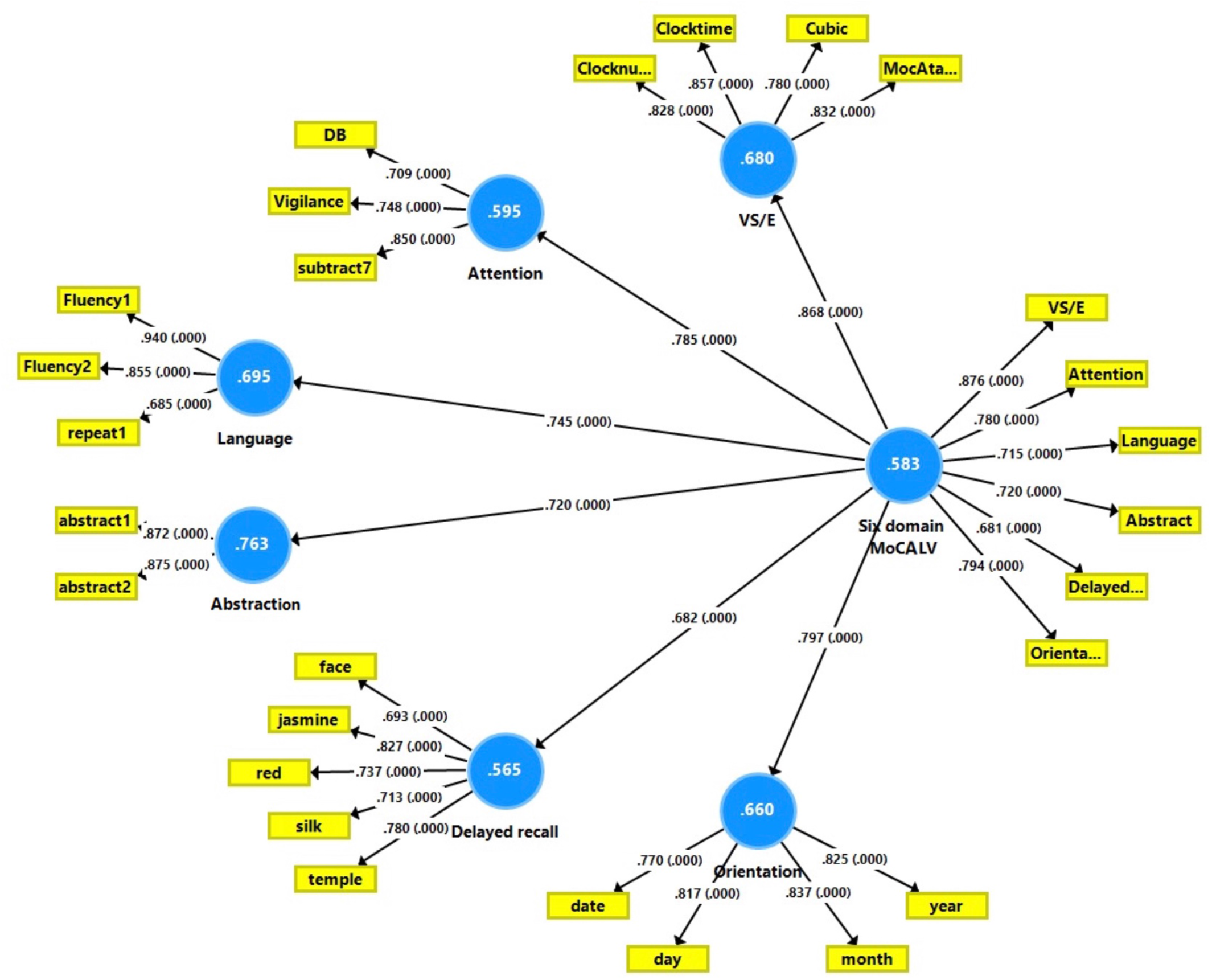
A second and reduced Partial Least Squares (PLS) model after feature reduction by deleting the Naming subdomain and five MoCA items from the analysis. A “six-domain MoCA” latent vector (LV) is constructed based on the scores on the six subdomains, which were entered as indicators. This LV was connected with all LVs extracted from the selected items of the six subdomain scores, dubbed the Visuospatial/Executive (VS/E), Attention, Language, Abstraction, Delayed Recall, and Orientation LVs. Shown are the loadings (with *p* values) for the outer models and path coefficients (exact *p* values) for the inner model are shown. White figures in blue circles denote the Average Variance Extracted (AVE).

### 3.4. Construction of the MoCA-Brief scale

Consequently, the model underwent removal of items that had the lowest correlation with their LV and retention of items that were the highest representative of the subdomains. First, we retrieved two items of each LV, namely Clock Time and Clock Number (VS/E), Vigilance and Subtract 7 (Attention), Fluency 1 and 2 (Language), Abstraction Pair 1 and 2 (Abstraction), Jasmine and Temple (Delayed Recall), and Month and Year (Orientation). Nevertheless, the convergence validity of the LV extracted from these 12 items was less adequate with AVE < 0.5 (0.45), although composite reliability (0.889), Cronbach’s alpha (0.750), and rho_A (0.752) were adequate. Moreover, six items showed loadings < 0.65 on this 12-item LV, namely Fluency 2 (indicating that the Fluency score is a better indicator than the dichotomized Fluency 2 indicator), Abstraction Pair 1 and 2, Jasmine, Temple, and Vigilance. Therefore, these six items were deleted from the outer model and the additional removal of Clock Number further improved convergent validity. **Figure 3** shows the final model which comprises five items, namely Clock Time, Fluency 1, Subtract 7, Month, and Year. This five-item solution showed adequate AVE (0.599), composite reliability (0.822), Cronbach’s alpha (0.832) and rho_A (0.833). The model quality was more than adequate with SRMR=0.033. All items loaded significantly (*p* < 0.001) and adequately (> 0.75) on this factor. **Figure 3** also shows that the LV extracted from these items is strongly associated with the total TMSE and MoCA score, as well as with the different subdomain scores. As such, we constructed a short version of the MoCA based on five items, which showed adequate convergent, construct, and concurrent validity, as indicated by the strong correlation with the total TMSE, total MoCA, and MoCA subdomain scores.

**Figure 3.**
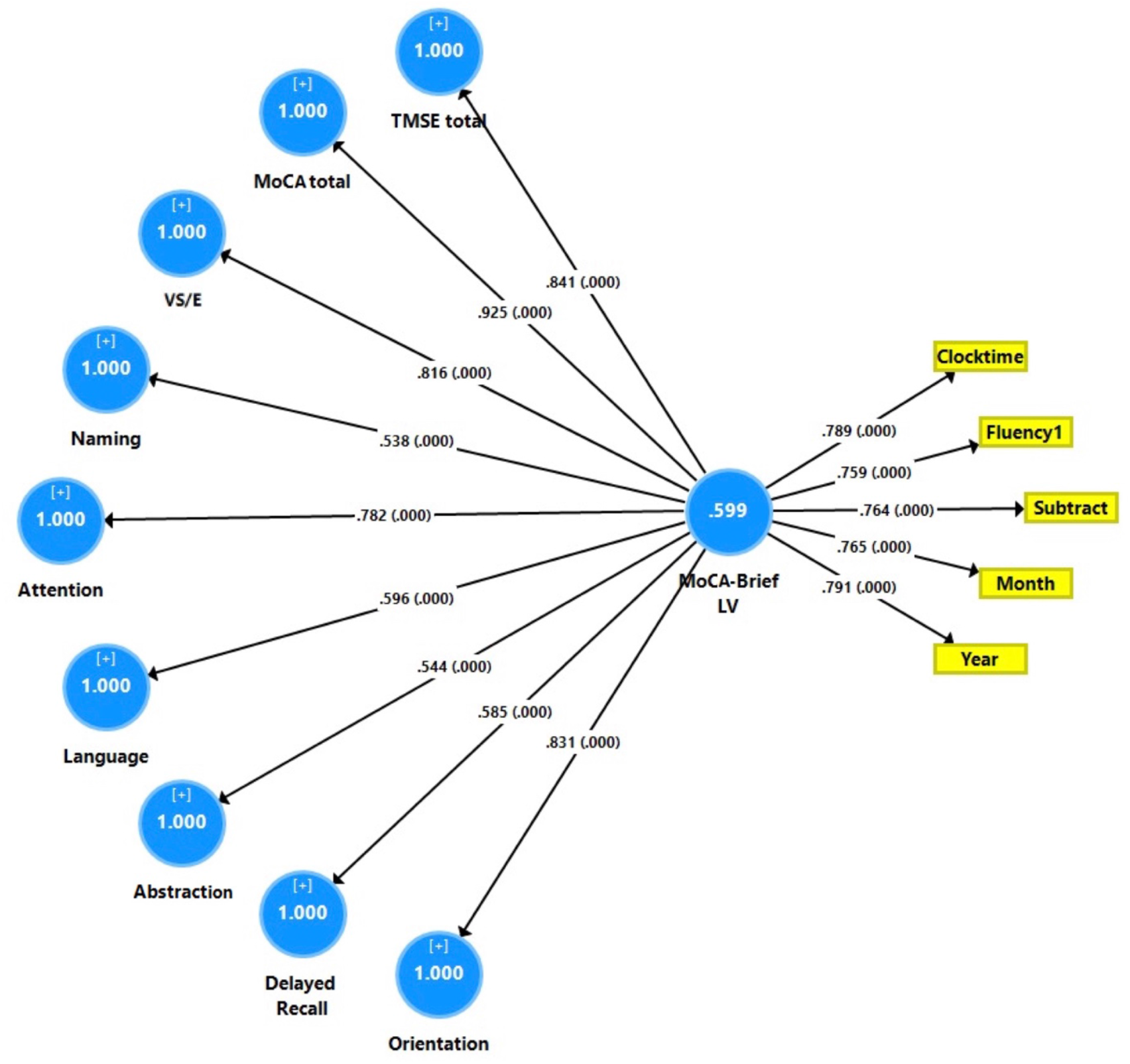
The Partial Least Squares (PLS) model showing the construct validity of the five-item MoCA-Brief. One latent vector (LV) was extracted from five selected MoCA items which were entered as indicators of the “five-item MoCA-Brief” LV. This LV was connected with the total Thai Mini-Mental State Examination (TMSE) and Montreal Cognitive Assessment (MoCA) scores and the seven subdomain scores of the MoCA. Shown are the loadings (with *p* values) for the outer models and path coefficients (exact *p* values) for the inner model are shown. White figures in blue circles denote the Average Variance Extracted (AVE).

Consequently, we checked whether the use of the dichotomized Fluency 2 indicator instead of the Fluency score resulted in adequate psychometric values. This model, which comprises Clock Time, Fluency 2, Subtract 7, Month, and Year, showed adequate AVE > 0.562, composite reliability > 0.864, Cronbach’s alpha (0.803) and rho_A (0.812). The model quality was adequate with SRMR = 0.037. All items loaded significantly (*p* < 0.001) and adequately (> 0.763), except Fluency 2 (0.616). Nevertheless, the latent variable scores computed for both the five-item models showed a correlation coefficient of 0.982. As such, to facilitate the use of the MoCA-Brief in clinical practice, the dichotomized Fluency index can be used to compute the sum of the five items. Nevertheless, for research purposes, it is more adequate to use the Fluency score together with the other four items.

### 3.5. Use of the MoCA-Brief as a diagnostic criterion for AD

The mean (standard deviation) values of the factor scores of the five-item MoCA in controls, aMCI, and AD are shown in **Table 1**. There were significant differences among the three groups and the factor scores decreased from controls to MCI and AD. **Figure 4** shows the mean values of the five-item factor score (with Fluency 1) and total MoCA sum (both in z transformations) in the three study groups. Results indicate that the distribution of the five-item factor score is nearly similar to that of the total MoCA score. We have also performed neural network analysis with the diagnosis AD and controls as output variables and the five items of the MoCA-Brief as input variables. The percent incorrect predictions were fairly constant between the training (5.8%), testing (0.0%), and holdout (3.1%) samples, whilst the sums of squares error were highly significantly decreased from the training (3.258) to the testing (0.011) conditions. The network was trained using two hidden layers with 4 units in layer 1 and 3 units in layer 2. Identity was the activation function in the output layer, whilst hyperbolic tangent was used in the first hidden layer. The partitioned confusion matrix showed an area under the ROC curve of 0.982 with an accuracy of 89.4% in the holdout sample with sensitivity = 85.7% and specificity = 92.3%. The relative importance of the input variables can be seen in **Figure 5**. Clock Time, Fluency 1, Subtract 7, and Month were the most important determinants of the model’s predictive power while Year followed at a distance.

**Figure 4.**
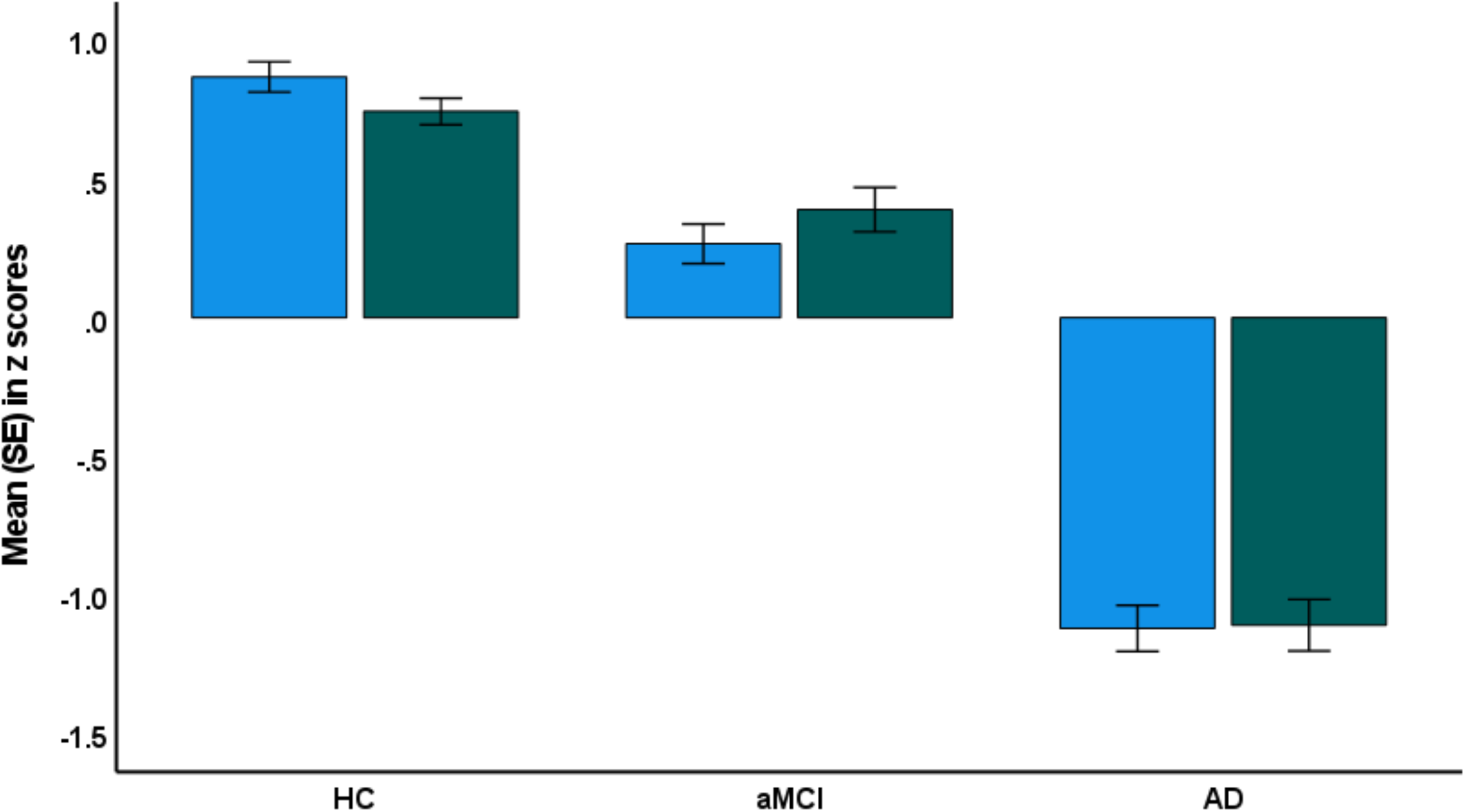
Mean (SE) values (in z scores) of the five-item MoCA-Brief factor score (green colour) and the total MoCA score (blue colour) in three study groups, namely healthy controls (HC), amnestic mild cognitive impairment (aMCI) and Alzheimer’s Disease (AD).

**Figure 5.**
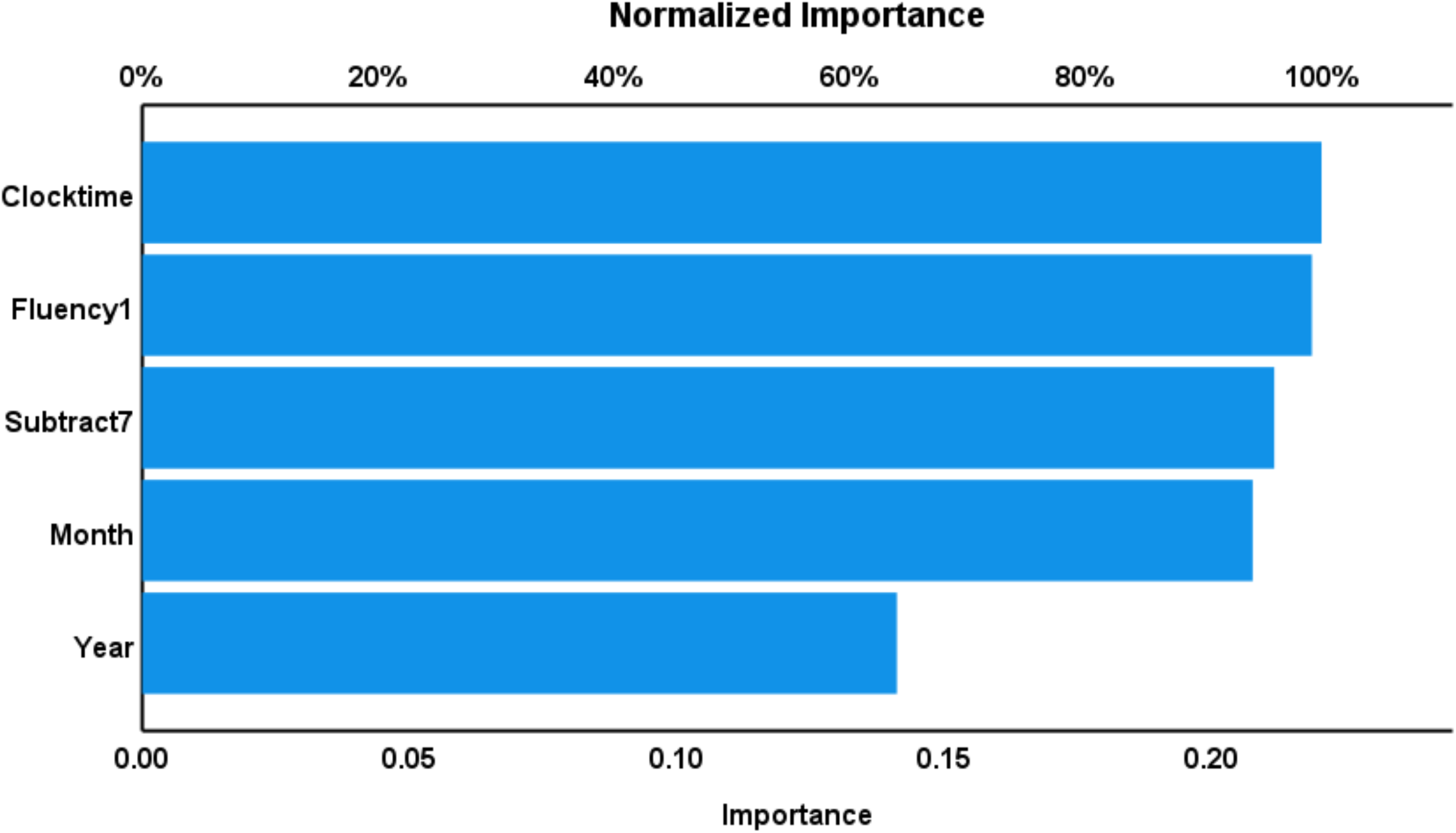
Results of neural networks analysis. This figure shows the relative and normalized importances of the five input MoCA-Brief items. Clock Time, Fluency 1, Subtract 7, and Month were the most important determinants of the model’s predictive power while Year followed at a distance.

### 3.6 Initial validation with the five-item MoCA

The test-retest and inter-rated reliability of the MoCA-Brief (based on the dichotomized Fluency indicator) was tested on 10 controls, 10 aMCI individuals, and 10 AD participants. Results show high test-retest reliability (*r* = 0.951, *p* < 0.01, and Cronbach’s alpha = 0.975) and inter-rater reliability (Kappa = 0.947 at *p* < 0.01).

## 4. Discussion

### 4.1 The construct validity of the MoCA subdomains is not optimal

The first major finding on this study is that not all items loaded highly onto their subdomains and that deleting some items improved the construct validity of the models. Although a general factor was present and the subdomains had high loadings on it, the observation that some items did not have satisfactory loadings on some subdomains implies that the total score on all items is not the most accurate assessment of cognitive state. It is difficult to compare these findings to the existing literature as, to the best of our knowledge, there is a lack of PLS analysis to evaluate the validity of the MoCA. Nevertheless, the finding that the full MoCA may not be entirely suitable is in line with some studies that used comparable methods. A study carrying out a cross-sectional analysis of data from the Irish Longitudinal Study on Ageing did not find an appropriate model for the MoCA and the cross-loading of items had no particular pattern, no general factor present, and many loadings were < 0.6 (Coen *et al*., 2016). These authors concluded that the MoCA may not be able to provide in-depth domain-specific neuropsychological assessment, but nevertheless was still a useful screening instrument. Similarly, discrepancies were also found between the EFA and CFA when the MoCA factor structure was examined in a large cohort of early PD patients (Smith *et al*., 2020). The alternative six-factor solution (1. Short-term recall, 2. Visuospatial-executive, 3. attention/working memory, 4. Verbal-exclusive, 5. Orientation, and 6. Expressive language) found from EFA in the early PD study was not supported by CFA. Collectively, there has been some indication that the MoCA does not show unidimensionality.

Contrastingly, other studies have reported that the MoCA contained a general factor, being unidimensional. Analysis of several models of the Portuguese MoCA using CFA found that a six-factor model (1. Executive functions, 2. Language, 3. Visuospatial skills, 4. Short-term memory, 5 Attention, concentration, and working memory, and 6. Temporal and spatial orientation), based on the conceptual model proposed by the original authors, had the best fit (Freitas, Simões, Marôco, *et al*., 2012). This six-factor model also had loadings ≥ 0.6 for all items except two: phonemic fluency in factor 1 (executive function) and factor 2 (language) at 0.21 and 0.55, respectively. Another comprehensive study on the psychometric properties of the MoCA in the Japanese population used a Rasch model to examine dimensionality and found evidence of multidimensionality (*p* < .010) (Sala *et al*., 2020). Upon further inspection, the authors reported that the MoCA was estimated to have eight factors (using parallel analysis) and contained a general factor (as suggested by the inspection of eigenvalues: first eigenvalue = 6.40 and second eigenvalue = 1.67). The author’s hierarchical EFA on the seven first-order factors that assumed a general factor, as suggested by the parallel analysis, found that all items loaded onto the general factor and the seven subfactors approximately corresponded to the original author’s seven subsets. The model estimated by the EFA analysis was confirmed by the CFA model. It is notable that when the authors in the study mentioned that all items loaded onto the general factor, most of the loadings were < 0.6, with only orientation year and day having loadings ≥ 0.6. This general factor is considered inadequate for the threshold in this present study.

The low loadings of Clock Circle, Lion, Repeat 2^nd^ Sentence, Digit Forward, and Place test items observed on their subdomains may be due to the observation that these items have low discriminatory power. In the Thai population, it has been observed that the Clock Circle, Lion, and Digit Forward task was generally too easy for patients, whilst the Repeat 2^nd^ Sentence task was too difficult. Furthermore, as the Place task required patients to name the hospital, this task was observed to be too difficult for elderly patients as they normally spend a significant amount of time at home and/or they may not have been hospital regulars.

After the removal of these five items, all remaining MoCA items now had loadings > 0.65 on their subdomains, and all six subdomain loadings on to the general factor remained high (all > 0.65). Therefore, the removal of these five MoCA items improved the assessment of cognitive deficits.

### 4.2 MoCA-Brief

The second major finding of this study is that we were able to construct a short version of the MoCA (the MoCA-Brief), which shows accurate construct, convergent, and concurrent validity, as well as test-retest and inter-rater variability. Many studies have also attempted to develop short variants of the MoCA in the past. A comparison study between seven short MoCA (s-MoCA) variants found that these versions all had acceptable levels of discrimination of MCI/dementia from normal controls (Liew, 2019). However, the authors state that the s-MoCA by Roalf *et al*. (2016) had comparable performance to the original MoCA even across different education levels. The s-MoCA by Roalf *et al*. (2016) was developed using item response theory and computerized adaptive testing simulation, resulting in an eight-item MoCA, consisting of 1) Visuospatial/Executive: Trails, 2) Visuospatial/Executive: Clock drawing, 3) Language: Naming (rhinoceros), 4) Attention: Serial 7s, 5) Language: Fluency, 6) Abstraction: Watch, 7) Delayed Recall, and 8) Orientation: Place. The items in the s-MoCA that are similar to our five-item MoCA are Clock, Subtract 7, and Fluency, helping confirm the use of the three items. The combination of the remaining items selected for our five-item MoCA (Month and Year) has been previously shown to be a sufficiently sensitive and specific indicator for dementia (O’Keeffe, Mukhtar and O’Keeffe, 2011), and may have also been absent in the s-MoCA by Roalf *et al*. (2016) due to cultural differences. In Thailand, most of the elderly reside with their family or closest relatives, living a sedentary lifestyle at home, thus being void of orientation tasks to stimulate them daily. Accordingly, the Month and Year task would hold relatively high discriminatory power as the Place, Date, and Day tasks in the Orientation domain would be too difficult for the elderly as they are incredibly specific and have a relatively finite number of answers.

In a newer study, two Chinese s-MoCAs were developed: 1) Four-item MoCA containing the Clock Drawing, Digit span, Serial subtraction, and Recall item, and 2) Six-item MoCA containing the Trail making, Clock Drawing, Digit span, Serial subtraction, Abstraction, and Recall item (Tan *et al*., 2021). From these two Chinese s-MoCA versions, a strong positive correlation between the short versions and the standard MoCA was found (Spearman r = 0.915 and 0.963, *p* < 0.001), and had a similar discriminatory performance for cognitive impairment to the full MoCA (sensitivity 0.91, specificity 0.82). Other s-MoCA versions have also been developed and tested in other populations including PD (Bezdicek *et al*., 2020) and stroke (Feng et al., 2021). The test items in the aforementioned MoCA short versions have some overlap with our MoCA-Brief.

Our five-item MoCA is initially a valid tool that should be used to screen for MCI and AD as it is significantly faster to administer and can still accurately capture the participant’s true cognitive state.

## 5. Conclusion

We demonstrate that the full MoCA does not have sufficient construct validity as five items have low loadings on the subdomain LV scores. We achieved adequate construct validity after the removal of those five items. Furthermore, we developed a MoCA-Brief scale comprising five items only, which shows adequate convergence and construct validity and concurrent, test-retest, and inter-rater validity. The MoCA-Brief constructed here awaits validation in independent samples and especially in other countries and cultures.

## Data Availability

The dataset and materials analyzed and/or generated during this study will be available from M.M. upon reasonable request once it has been fully utilized by the authors.

## 6. Acknowledgements

We would like to thank Professor Ziad Nasreddine for giving us the opportunity to carry out this research and Professor Jeffrey Cummings for his mentorship.

## 7. Statement of Ethics

Our research involved human participants. This study obtained ethical approval by the Institutional Review Board of Chulalongkorn University (Bangkok), Chao Phraya Abhaibhubejhr Hospital (Prachinburi), and Angthong Hospital (Ang Thong), Thailand, and is in accordance with the principles embodied in the Declaration of Helsinki. Written informed consent was provided by all participants and guardians of aMCI and AD patients prior to participation in the study.

